# Healthfulness of the Food Environment in Putrajaya, Malaysia: Implications for Public Health Action

**DOI:** 10.1101/2025.11.16.25340357

**Authors:** Nur Farhana Mohammed Noor, Siti Aisah Mokhtar, Halimatus Sakdiah Minhat, Siti Anom Ahmad, Zarirah Adzni Mokhtar

## Abstract

**Introduction:** The prevalence of overweight and obesity in Malaysia has reached unprecedented level, with Putrajaya recording among the highest rates. Despite this, limited attention has been given to understanding the local food environment, which plays a crucial role in shaping dietary behaviours. This observational study aimed to assess the healthfulness of food establishments in Putrajaya and provide insights into their implications for public health.

**Methods:** A total of 745 food establishments, including 80 food stores and 665 eateries, were systematically identified, geocoded using Google Maps, and validated through cross-referencing with supplementary sources. Each establishment was assessed using the Nutrition Environment Measures in Stores (NEMS-S) and Nutrition Environment Measures in Restaurants (NEMS-R) instruments. capturing the availability, quality, pricing, and promotion of healthier food options. NEMS scores were computed and compared across establishment types. Descriptive statistics and spatial mapping were used to summarize and illustrate patterns of food environment healthfulness.

**Results:** Healthy food items such as whole-wheat bread, low-sugar cereal, and healthier beverages were more commonly available, whereas fruits and vegetables remained limited across stores. Supermarkets recorded the highest overall NEMS-S scores, followed by fresh markets and grocery shops, while convenience stores performed poorest. Among eateries, healthier menu options such as vegetable dishes, healthy entrées, and 100% fruit juice were more readily accessible as compared to fruits, whole-grain bread, and brown rice. 24-hour restaurants scored the highest NEMS-R scores, whereas fast-food outlets presented the greatest barriers to healthy eating. Spatial analysis revealed pronounced disparities, with healthier outlets clustering in northern precincts, while less healthy environments were more widely dispersed throughout the city.

**Conclusion:** The study highlights notable disparities in the availability of healthy food options across Putrajaya. Strengthening policies and initiatives that foster healthier retail and dining environments is essential to promote nutritious eating and mitigate obesity risks in this planned urban community.

## 1.0 INTRODUCTION

The food environment refers to the context in which individuals purchase and consume food, encompassing various dimensions such as food availability, accessibility, and affordability that collectively shape dietary behaviours and health outcomes [1]. Therefore, the food environment serves as a critical link between the broader food system and individual preferences and dietary norms [2]. The food environment can be influenced by a dynamic interplay of factors, including physical factors such as proximity and density of food outlets, economic elements such as pricing and income levels, policy regulations around food safety and marketing, as well as the sociocultural influences that affect individual preferences, traditions, and dietary norms [1]. Together, all these elements play a role in determining the type of food options that are easily accessible, affordable, and acceptable in the local community, which ultimately shaping the foods that individuals are most likely to consume.

Recently, with the increased prevalence of non-communicable diseases (NCDs), particularly obesity, diabetes, hypertension, and other diet-related diseases, more attention has been shifted towards the food environment due to its critical role in influencing public health [3]. Communities living with a high density of fast-food outlets, convenience stores, and take-out restaurants have been shown to report higher rates of obesity and related health problems [4–9]. These food environments predominantly offer energy-dense and nutrient-poor options that promote unhealthy dietary intake. The easy access to inexpensive, calorie-dense foods, coupled with a scarcity of healthier alternatives, has led to what is known as a ‘food swamp’, an area with an overwhelming availability of unhealthy food options. Similarly, areas with restricted access to affordable, nutritious foods such as fresh fruits and vegetables, have created so-called ‘food deserts’, which further constraint residents’ ability to make healthy dietary choices [10]. Both food swamps and food deserts, which are considered obesogenic environments, are associated with poor dietary intake and increased obesity prevalence, largely driven by the widespread availability of processed and high-calorie foods [5–6,11].

According to the Theory of Planned Behaviour, human behaviour can be shaped by the complex interplay between personal, behavioural, and environmental factors. In the context of the healthy food environment, the theory delineates how improved availability, accessibility, and affordability of certain food options can foster positive attitudes towards these foods, subsequently normalise their consumption, and create social norms that favour healthy eating habits. More importantly, individuals will foster a sense of control over their ability to make healthy food choices [12]. On the other hand, living in an obesogenic environment with readily available unhealthy options such as fast food and sugary snacks may lead individuals to choose these foods largely due to the convenience and accessibility they offer.

Nevertheless, mirroring global trends, obesity is a significant public health concern in Malaysia. According to the National Health Morbidity Survey (NHMS) 2023, more than half (52.9%) of the adult population in Malaysia is either overweight or obese, putting Malaysia as one of the countries with the highest prevalence of obesity in Southeast Asia. Putrajaya, the federal administrative capital of Malaysia, closely mirrors the national trend, with over 50% of the adult population classified as overweight or obese [13]. Despite advantages such as high levels of health literacy and a built environment that supports physical activity in Putrajaya, the persistently high prevalence of obesity points to the local food environment that influences dietary habits as a possible contributing factor. Therefore, this study aimed to assess the healthfulness of food establishments in Putrajaya and provide insights into their implications for public health.

## 2.0 METHOD

This observational study was conducted in Putrajaya, the federal administrative capital of Malaysia. Established in 1995, Putrajaya was designed according to the ‘garden city’ concept, with 38% of its land area allocated to green and open spaces. The city layout balances government offices, residential and commercial areas, parks, open spaces, and protected wetlands. Home to over 30,000 residents, most of whom are government employees under housing subsidies, Putrajaya presents a unique urban and demographic profile. Paradoxically, despite its planned infrastructure and abundant green spaces, Putrajaya has the highest prevalence of overweight and obesity in Malaysia.

To assess the diverse food environment in the region, the study employed two validated instruments: the Nutrition Environment Measures Survey for Supermarkets (NEMS-S) [14] and for Restaurants (NEMS-R) [15]. Both tools were used to evaluate the availability, accessibility, and quality of healthy food options in Putrajaya.

### 2.1 Data preparation

The geographical coordinates of food establishments including food stores and eateries in Putrajaya were identified and geocoded from online sources, namely Google Maps. Following that, the data were validated through cross-referencing with additional sources, including business directories, meal delivery service websites, relevant social media platforms, and on-site verification. This extra step was necessary to ensure the accuracy and completeness of the data. Subsequently, all food establishments were then classified into two categories: (1) Food stores: Supermarkets, grocery stores, convenience stores, and fresh markets; and (2) Eateries: Restaurants, 24-hour restaurants, coffee shops, bakeries, fast-food outlets, and hawker stalls.

Next, each identified food store and eatery was systematically audited using the NEMS-S and NEMS-R tools, respectively, to assess their contribution to the local food environment. The scores were computed by analysing food items offered on the menus at these-food establishments. In addition, the nutritional information on food packaging was also evaluated based on the Dietary Guidelines of Malaysia [16]. The assessment tool was adapted to better reflect local cuisine. For example, the salad options were enhanced to include cooked vegetables, and brown rice was incorporated as an alternative to chips. Similar adjustments were previously made by Cai [17] when researching Asian cuisine, with the adapted tool demonstrating high reliability and validity.

**Figure 1.**
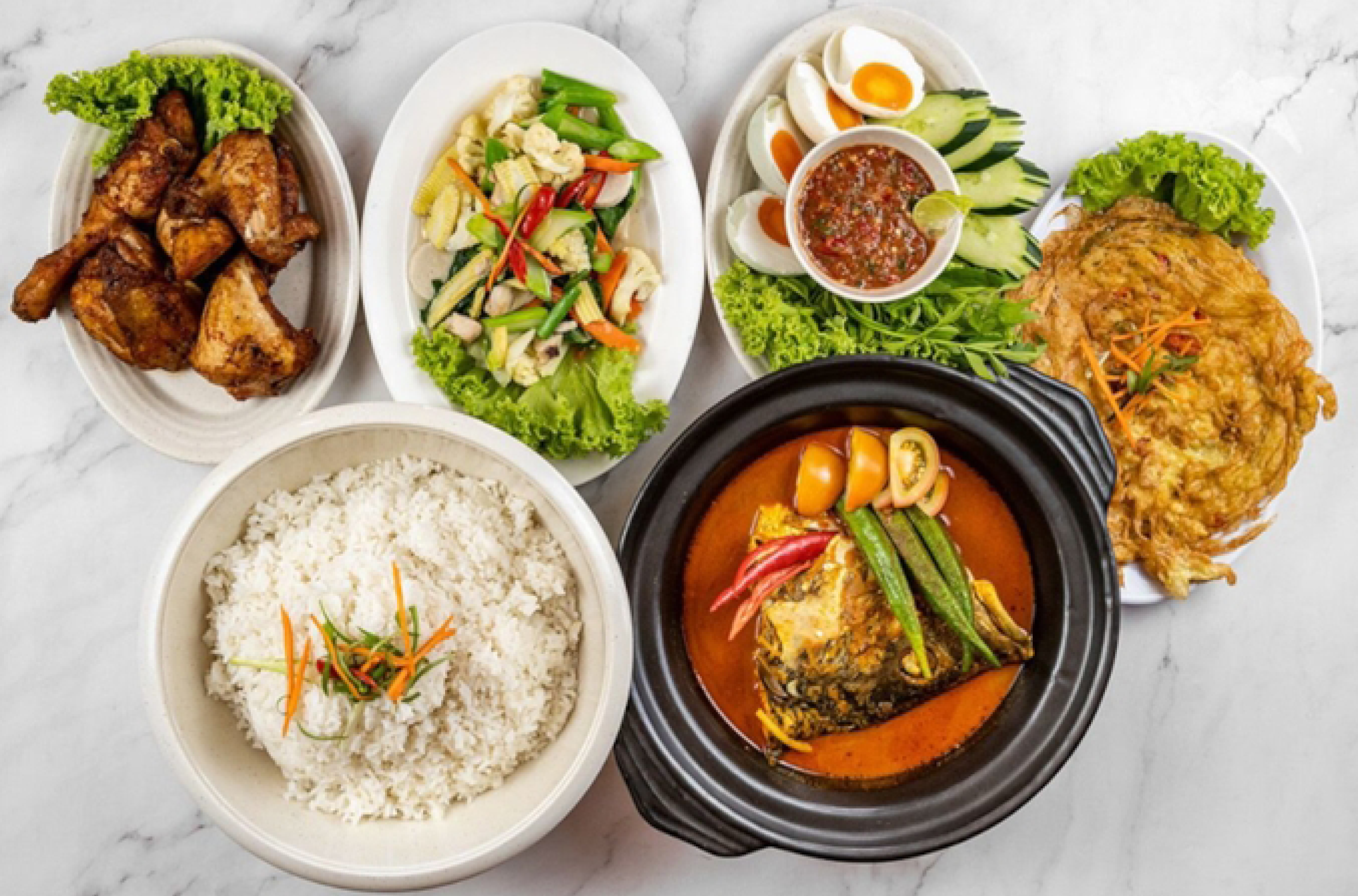
Example of a local meal; white rice served with choices of protein and vegetables as side dishes.

To minimize potential bias, two independent raters conducted the assessments simultaneously. Both raters maintained discretion to avoid influencing retailer behaviour during the observation process. All food item observations were independently documented, cleaned, and entered into two distinct databases. Data quality assurance involved cross-validation of the two datasets, whereby any discrepancies were reconciled through averaging or consensus discussion. Inter-rater reliability analysis indicated a 99% agreement, confirming a high level of consistency between raters’ assessments.

### 2.2 Study instrument

NEMS-S and NEMS-R were adapted to evaluate the food environment of Putrajaya in this study. Both instruments have demonstrated high reliability and validity with Kappa values for inter-rater and intra-rater reliability above 0.80 and 0.72, respectively in previous studies [14,15].

NEMS-S assesses the healthfulness of food available in food stores, measuring key elements such as availability (score range from 0 to 30), price (score range from -9 to 18), and quality (score range from 0 to 6) of ten food items that are highly recommended for a nutritious diet. These items include a variety of fresh fruits and vegetables, skim or low-fat milk instead of whole milk, lean ground beef instead of regular ground beef, low-fat hot dogs instead of regular hot dogs, reduced-calorie frozen dinners instead of regular frozen dinners, diet or low-calorie soda instead of regular soda, 100% juice instead of juice drinks, lower-fat baked goods instead of regular baked goods, 100% whole grain bread instead of refined bread, and baked or low-fat snack chips instead of regular snack chips [14]. The total NEMS-S score is calculated by summing the availability, quality, and price components, with a total score ranging from -8 to 54, whereby higher scores indicate healthier food environments.

On a similar note, the NEMS-R is an observational tool used to assess the healthfulness of the food environment in restaurants. The NEMS-R scoring includes several components such as the availability of healthy meal options (e.g. entrées, salads, fruits, non-fried vegetables, whole grain bread, baked chips/ brown rice, and low-calorie/ low-fat beverages) with a score from 0 to 15. In addition, the tool also assesses the facilitators of healthy eating (score ranging from 0 to 8), barriers to healthful eating (score ranging from -5 to 0), and the presence of a healthy kid’s menu (score ranging from -3 to 9). The total score of the NEMS-R ranges from - 5 to 23, whereby higher scores indicate healthier food environments. A meal is classified as healthy if it meets predetermined nutritional criteria. For this study, the “healthy kid’s menu” category was omitted from the assessment.

### 2.3 Data analysis

For each tool, individual item scores were aggregated to produce subscale scores, and the total score for each food establishment was obtained by summing the subscale scores across all categories. Descriptive statistics, including means and standard deviations, were computed to summarize NEMS scores across outlets.

Subsequently, each premise was geocoded and mapped in ArcGIS Pro (Esri Inc., Redlands, CA, USA) to visualize the spatial distribution of healthy and unhealthy food establishments across Putrajaya. Kernel Density Estimation (KDE), implemented using the Spatial Analyst tool in ArcGIS, was employed to identify areas of high concentration of healthy and unhealthy food environments, enabling detailed analysis of spatial clustering patterns.

This study was conducted with approval from the Ethics Committee for Research Involving Human Subjects Universiti Putra Malaysia (JKEUPM-2024-354). Data collection involved publicly accessible food establishments. No protected or privately restricted lands were accessed during the study.

## 3.0 RESULTS

A total of 99 geocoded food stores were initially identified. However, 19 were excluded due to geocoding duplication (4), located in private compounds (6), classification as home-based enterprises (3), temporarily ceased operations (2), or permanently closed (4). Among the 80 food stores in the final analysis, four were supermarkets, 36 were grocery stores, 30 were convenience stores, and 10 were fresh markets, including wet markets, butcher shops, and greengrocers.

Among these stores, items such as whole-wheat bread (80.5%), diet soda or 100% juice box (79.3%), low-fat milk (77.9%), low-sugar cereal (71.4%), and low-fat hotdogs (71.4%) were among the more readily available items across the stores. In contrast, vegetables (50.7%) and fruits (41.6%) were less commonly available in food stores. The most frequently stocked vegetables were choy sum (sawi), tomato, and cabbage while the most common fruits were oranges and bananas, followed closely by apples and pineapples. Overall, some healthy food items were less common in these stores, such as low-fat frozen dinners (44.2%), low-fat baked products (6.5%), lean meat (5.2%), and low-fat chips (2.6%). The percentage of food stores offering healthy food items is summarised in Figure 2.

**Figure 2.**
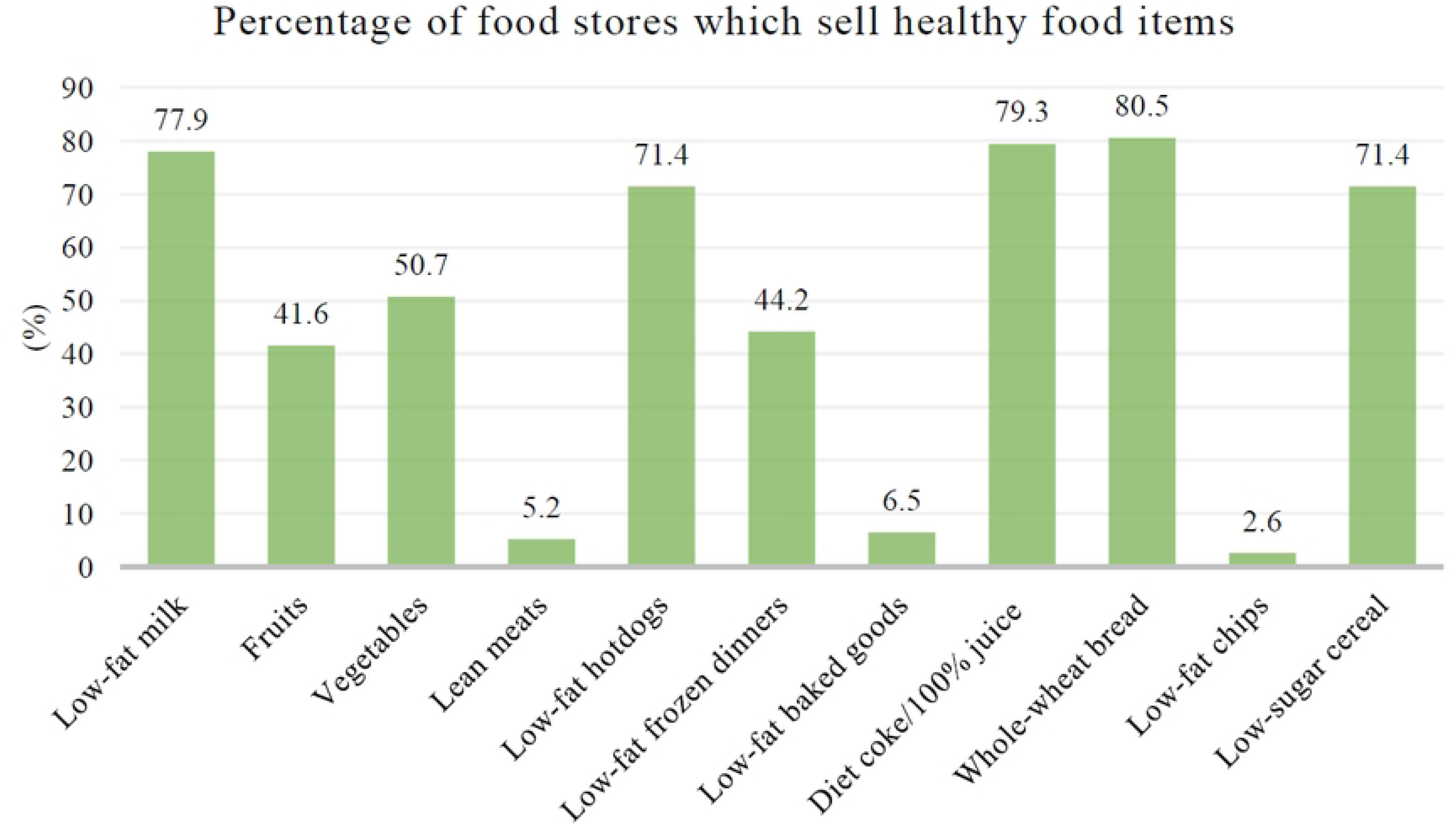
Percentage of food stores in Putrajaya which sell healthy

Additionally, Table 1 shows the availability of healthy food items according to the types of food stores. All supermarkets, as well as most convenience stores and grocery stores, offered low-fat milk, diet soda, 100% juice, low-fat hotdogs, whole-wheat bread, and low-sugar cereal. Supermarkets, fresh markets, and grocery stores were the primary locations where fruits and vegetables were sold. In addition, some stores carried low-fat frozen dinners. However, lean meat was less commonly available. Only several stores provide low-fat meat cuts but they did so without any nutritional labelling.

**Table 1.** Percentage of healthy food items sold by each type of food stores.

| Type of food | Supermarket (%) | Grocery store (%) | Convenience store (%) | Fresh market (%) |
| --- | --- | --- | --- | --- |
| Low-fat milk | 100 | 80.6 | 86.7 | 25.0 |
| Fruits | 100 | 55.6 | 16.7 | 50.0 |
| Vegetables | 100 | 69.5 | 16.7 | 75.0 |
| Lean meats | 25.0 | 2.8 | 0 | 37.5 |
| Low-fat hotdogs | 100 | 63.9 | 83.3 | 50.0 |
| Low-fat frozen dinners | 75.0 | 13.9 | 80.0 | 75.0 |
| Low-fat baked goods | 100 | 2.8 | 13.3 | 50.0 |
| Diet soda/100% juice | 100 | 80.5 | 93.3 | 50.0 |
| Whole-wheat bread | 50.0 | 77.8 | 100 | 12.5 |
| Low-fat chips | 50.0 | 19.4 | 0 | 0 |
| Low sugar cereal | 100 | 58.3 | 100 | 12.5 |

Across different types of food stores, supermarkets recorded the highest average total score (28.23), as well as scores for availability (19.3) and quality of healthy food (5.8). Fresh market and grocery shop came in second and third, with total average scores of 16.8 and 15.8, respectively. In comparison, convenience stores had the lowest total average score (15.5) and quality score (0.7) despite achieving the highest average score for price (4.17) (Table 2).

**Table 2.**
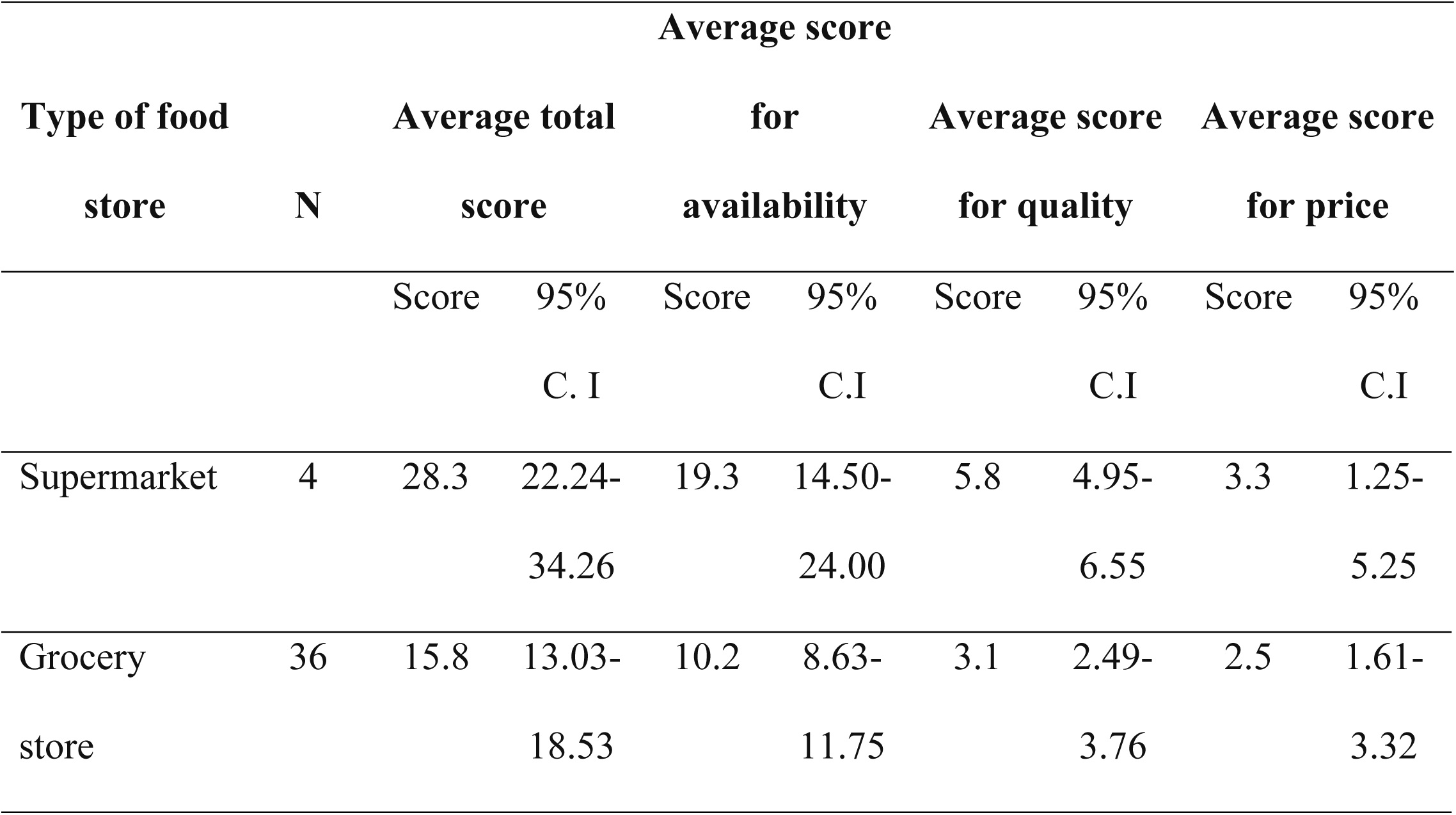
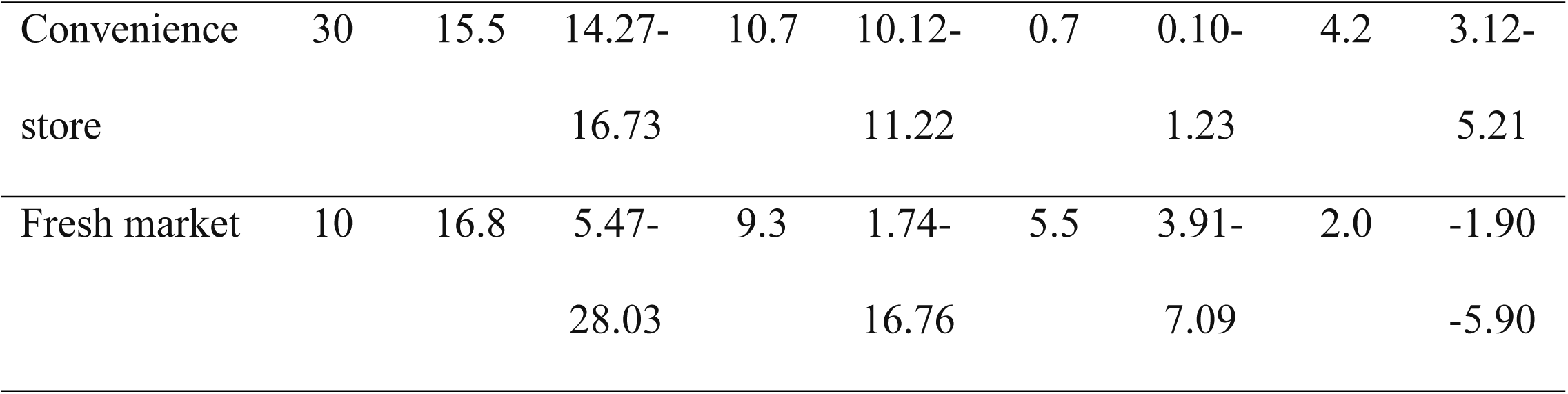
Overall score and the availability, quality, and pricing sub-scores for each category of food stores.

Next, a total of 665 food eateries were included in this study, comprising 404 restaurants that operated during normal opening hours, 13 restaurants that operated 24 hours, 74 coffee shops, 27 bakeries, 110 hawker stalls, and 37 fast-food outlets. Overall, only a few dining places offered healthy food options on their menus. Among the healthy items available, 25.4% of eateries offered vegetables as the main dish or a side with rice, 22.3% offered non-fried vegetables, and 21.9% provided healthy entrée or soup. Approximately 19.5% of eateries offered 100% juice, 7.4% served low-fat salad dressings, 4.5% included fruits, 1.4% offered whole-grain bread, 1.1% offered brown rice, and 0.5% included low-fat milk options on their menu (Figure 3).

**Figure 3.**
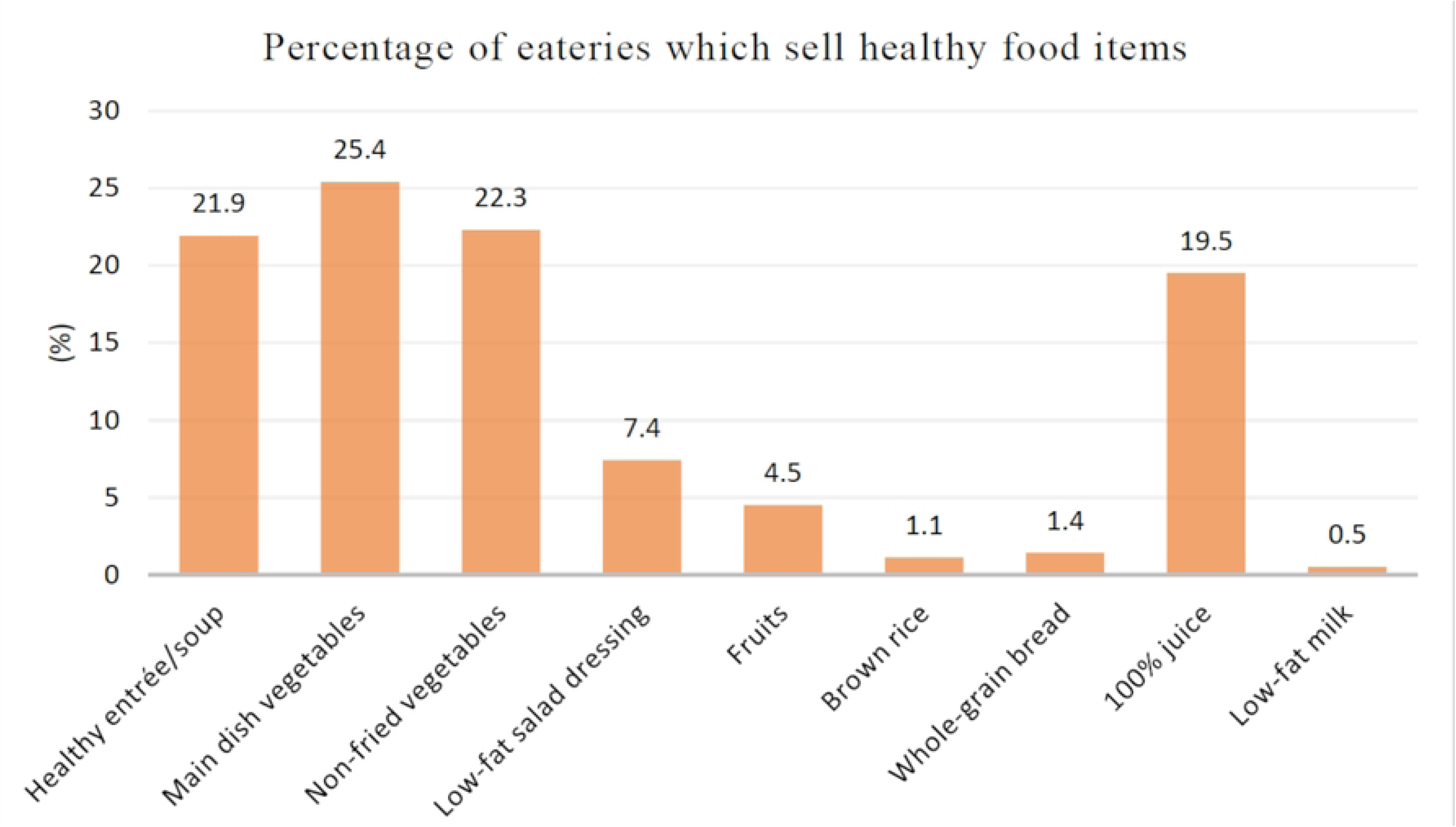
Percentage of eateries in Putrajaya that sell healthy food items

Table 3 shows the percentages of healthy food options available across each type of eatery in Putrajaya. Notably, restaurants served all the listed healthy food items. Numerous 24-hour restaurants, commonly known as ‘*mamak*’ restaurants, included healthy entrée or soup, main-dish vegetables, non-fried vegetables, whole-grain bread, and 100% juice on their menus. Additionally, most coffee shops that served Western-style cuisines would include healthy entrée or soup, main-dish vegetables, salad, fruits, and 100% juice on their menus. In comparison, most hawker stalls primarily offered snacks and fast food, with only a few featuring healthy entrée or soup, main-dish vegetables, and non-fried vegetables. Among fast-food outlets, some offered healthy entrée or soup, main-dish vegetables, and whole-grain bread.

**Table 3.**
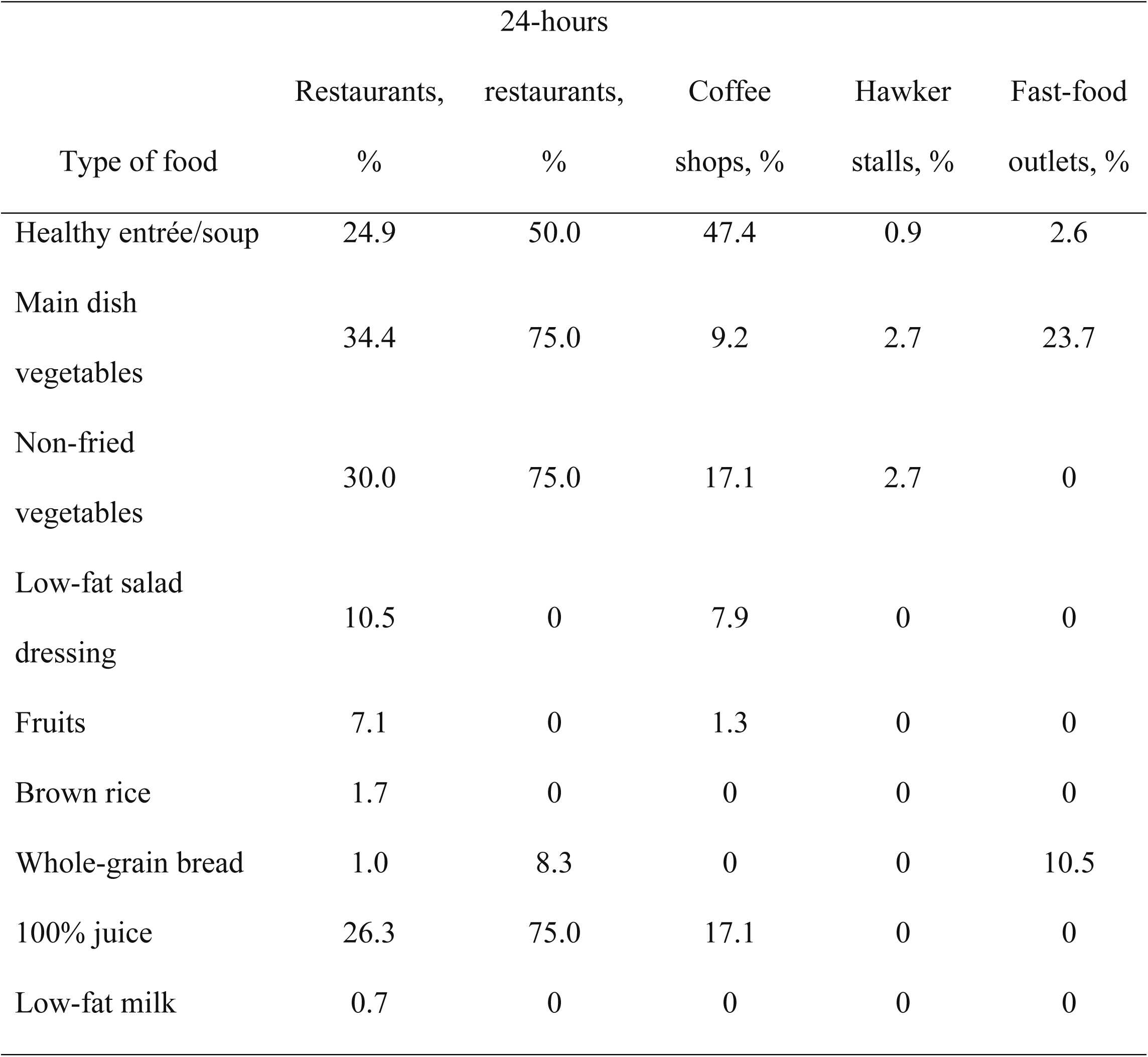
Percentage of healthy food items sold by each type of eateries.

A comparison of scores across the food eateries revealed that 24-hour restaurants recorded the highest average total score (4.2), as well as the score for the availability of healthy eating options (4.2). Restaurants ranked second with a total average score of 1.8. These normal-hour restaurants also recorded the highest score for being a facilitator of healthy eating (0.04). Coffee shops, fast-food outlets, and hawker stalls recorded total scores of 1.2, 0.4, and 0.06 respectively. Fast-food outlets scored the highest in the category of “barrier to healthy eating” (0.8) while bakeries did not score any points in any category of healthy dining (Table 4).

**Table 4.**
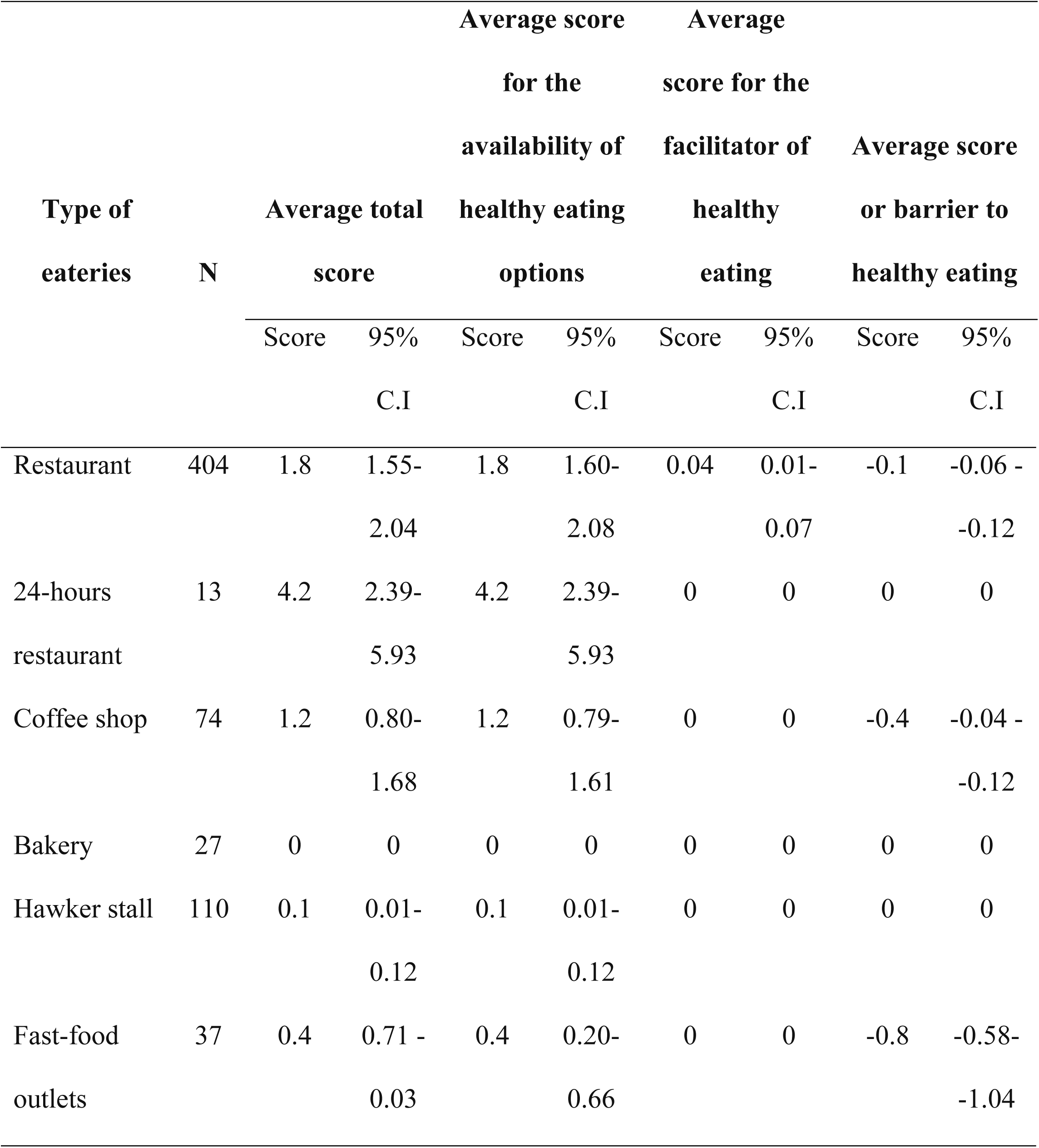
Average total score and sub-scores for each category of availability, facilitators, and barriers to healthy eating of food eateries in Putrajaya.

Subsequently, GIS-based map was generated for various types of food establishments, based on the NEMS-R and NEMS-S scores, to provide a detailed representation of the food environment in Putrajaya, as shown in Figure 4. In the map, square symbols denote the locations of food stores, while dot symbols represent eateries. The size of each symbol corresponds to the total NEMS score, with smaller symbols indicating lower scores.

**Figure 4.**
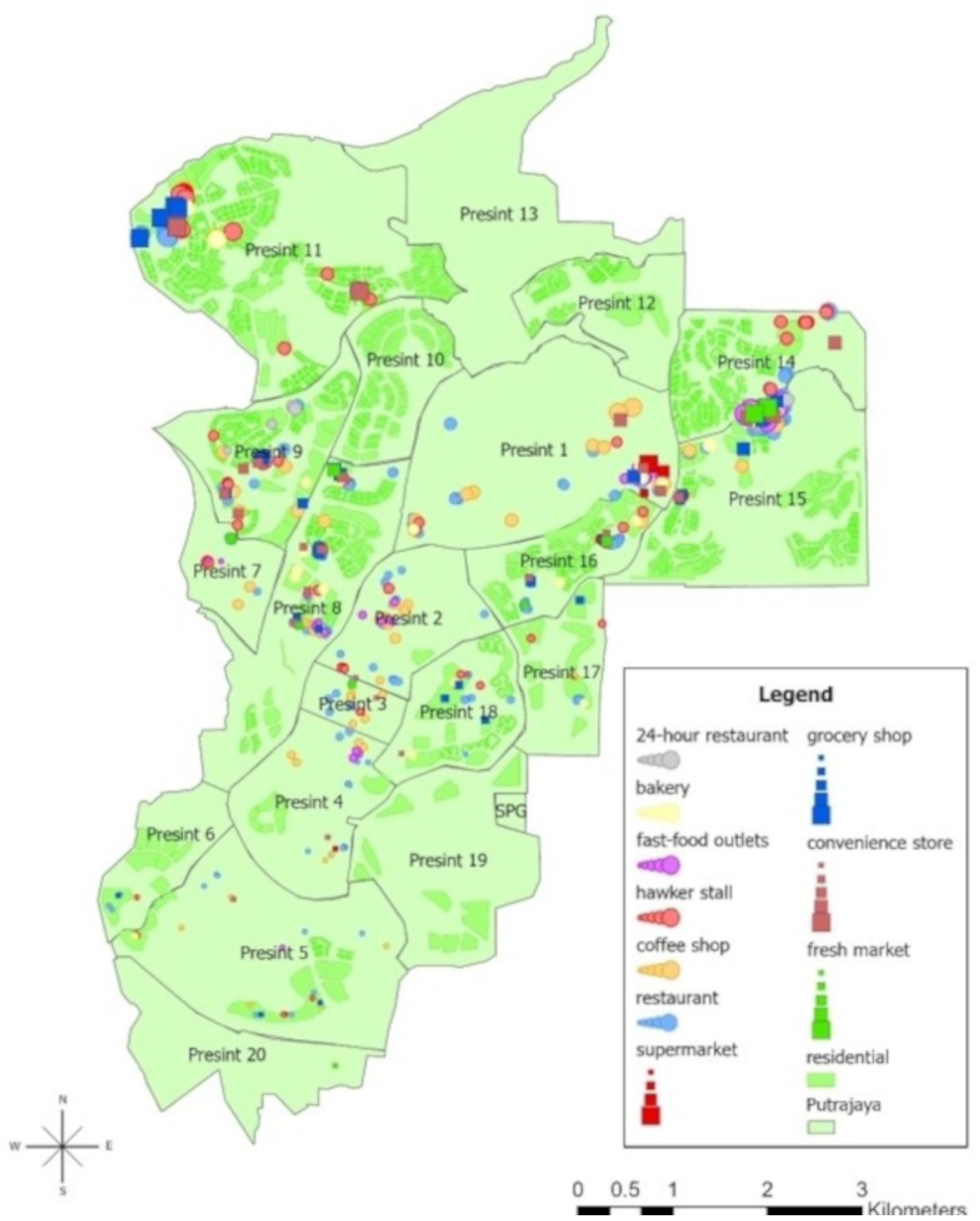
Spatial distribution of food stores and eateries in Putrajaya; squares denote the location of food stores while dots show the location of the eateries. The size of the dot/square was based on the total score, with a smaller size indicating a lower score.

Figure 4 illustrates the spatial distribution of food establishments (food stores and eateries) in Putrajaya. The proportional point in spatial data represented the NEMS-S and NEMS-R scores of food stores and eateries, with the size adjusted according to total scores. The smaller the symbols, the lower the scores.

Most food establishments are situated in shopping malls and commercial zones, with some located in office and residential areas. Overall, food establishments are unevenly distributed, with more concentrated in the northern areas of Putrajaya, particularly in Precincts 1, 2, 9, 11, 14, 15, and 16, whereby food establishments were in the form of clusters. In contrast, food establishments were more dispersed across Precincts 3, 4, 6, 17, and 18 in the southern part.

Convenience stores and grocery shops are found across most precincts but are most concentrated in Precincts 8, 9, and 15. Supermarkets and fresh markets are fewer and primarily located in Precincts 1 and 15, which function as major service and commercial hubs. Among eateries, restaurants and coffee shops are widespread, especially in Precincts 1, 2, 3, 4, 9, and 15, whereas fast-food outlets and bakeries are concentrated in Precincts 1, 8, and 15. Hawker stalls and 24-hour restaurants occur mainly in Precincts 8, 9, and 15.

When considering the healthfulness of the food environment, larger symbols representing higher NEMS scores, indicating healthier food options, are predominantly located in the northern part of Putrajaya, particularly in Precinct 1, 11, 14, and 15. In contrast, smaller symbols corresponding to lower NEMS scores are more widespread, particularly observed in the central and southern precincts, such as Precinct 2, 3, 4, 5, 8, 9, and 18 reflecting spatial disparities in the distribution of healthy food access across the city.

Furthermore, a GIS map was developed, as shown in Figure 5, to illustrate the spatial density of healthy and unhealthy food environments in Putrajaya, based on the NEMS-R and NEMS-S scores.

**Figure 5.**
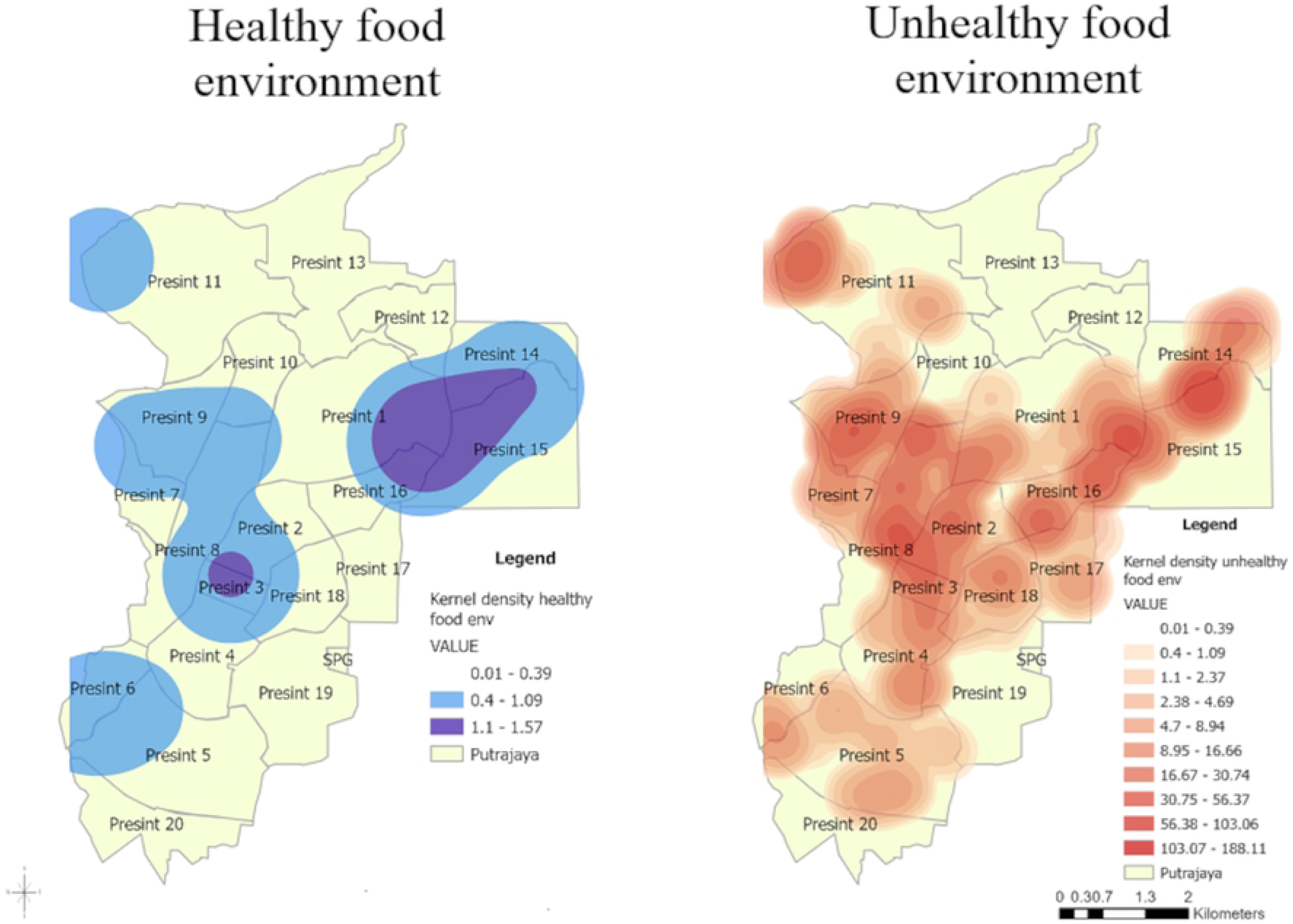
The density of healthy and unhealthy food environments in Putrajaya.

Figure 5 further delineates the density of healthy and unhealthy food environments in Putrajaya. Dark blue areas denote locations with the highest density of healthy food environments, with colours gradually lightening as the density decreases. The lightest blue colour indicates regions with the lowest density of healthy food environments. Contrarily, dark red areas represent the highest density of unhealthy food environments, with the colour becoming lighter as the density diminishes. Light red areas show the lowest density of unhealthy food environments.

Spatially, the healthy food environment demonstrates a more localized and concentrated pattern, primarily clustered in Precincts 1, 14, 15, and 16. In comparison, the unhealthy food environment exhibits a broader and more dispersed spatial coverage, with multiple clusters are evident throughout Putrajaya, particularly in Precincts 1, 2, 3, 4, 7, 8, 9, 14, and 15. Here, it is evident that the extent and intensity of the unhealthy food environment surpass that of the healthy food environment, indicating a greater availability and accessibility of energy-dense, nutrient-poor food outlets throughout the city.

## 4.0 DISCUSSION

The findings of this study shed light on important details about the food environment in Putrajaya, including the availability, accessibility, and quality of healthy food options for the residents. The availability of healthy food options is the primary focus of this study as it shapes the local food landscapes. In this study, supermarkets emerged as the leading source of nutrient-dense foods, offering a broader selection of fresh produce and healthy frozen items compared to other food retailers. Similar findings were reported in previous research [18]. Typically located in urban centres, supermarkets cater to a broad market and offer a sizable number of imported products, emphasising product quality, freshness, and perceived price value that aim at satisfying consumers’ needs [19,20].

On the contrary, grocery stores are usually small family-operated businesses situated in neighbourhood areas. They provide convenient access for nearby residents to purchase small quantities of grocery goods when the need arises [19]. Therefore, grocery store owners often prioritise daily essentials that are either likely to sell quickly or have a longer shelf life, such as processed and packed foods [21]. Unlike supermarkets with established distribution chains, the high costs associated with sourcing and preserving fresh produce may limit the range of healthy items offered by small grocery stores, as Malaysian consumers often view subpar food as unsafe [22–23]. Consequently, while grocery stores contribute to improved food accessibility in Putrajaya, they also inadvertently promote a less nutritious diet among the residents.

In contrast to previous studies [24,25], this study found that convenience stores contributed more favourably to the food environment than grocery stores. In recent years, the mushrooming of large chains of convenience stores in Malaysia has enabled these stores to provide a variety of pre-packed and ready-to-eat foods, including salads, healthy snacks, cut fruits, and frozen meals [24,26]. Furthermore, strong supplier relationships, strategic marketing distribution networks, and bulk-purchasing capacity allow them to stock a broader selection of healthier food options [19]. The ‘on-the-go’ or ‘grab-and-go’ business models of these convenience stores align well with the fast-paced lifestyle of many Malaysians, especially the working population in Putrajaya who prefer convenient and healthy food options [19,24,27]

Next, fresh markets that primarily sell produce such as poultry, seafood, fruits, and vegetables scored low in terms of the overall healthy food environment. This is likely due to the limited offering of nutrient-dense items like dairy, plant-based goods, and grains. Despite the lower overall scores, fresh markets play a crucial role in promoting healthy eating by providing easy access to protein, fruits, and vegetables, all of which are essential components of a well-balanced diet. Thus, their roles in safeguarding accessibility to fresh produce are invaluable in fostering healthy eating habits and supporting the local economies [28].

In terms of food eateries, the study found that most served nutritionally unbalanced meals, as previously documented, posing a significant barrier to healthy eating and potentially contributing to diet-related health risks [12,29]. Often, cost and profitability may influence the availability of healthy menu options in food eateries, as healthier ingredients like fresh produce, lean meats, and whole grains often cost more than processed alternatives. As a result, restaurateurs may resort to less costly ingredients to maintain competitive pricing and attract customers [30]. Furthermore, low demand for healthy food such as fruits and vegetables may have led many local retailers to prioritise more popular but less nutritious food items [2,31–32] Studies show that Malaysian diners highly value taste, freshness, appearance, price, and food safety [33–36] However, these demands always come at a higher price, hence becoming a potential barrier to healthy eating among individuals who frequently dine out [37–38].

Interestingly, 24-hour restaurants scored higher for healthfulness in this study compared to other eateries, which is unexpected given their typically high-calorie menu offerings. This result, however, may be explained by the presence of a few healthier options, such as vegetable dishes and 100% fruit juice, that are available in 24-hour restaurants but often absent from the menus of other dining establishments.

### 4.1 Policy implications

Overall, this study highlights a disparity in the distribution of healthy and unhealthy food options in Putrajaya, reflecting an uneven food environment that may influence dietary behaviors and contribute to the high prevalence of overweight and obesity. Additional research and targeted interventions are needed to address this imbalance and promote healthier food environments for the community. A comprehensive strategy encompassing expanded availability of nutrient-dense food, removal of cost barriers, and increased public awareness is necessary to improve the food environment. Currently, small retailers like grocery stores account for the majority of retail sales of food items (56%), followed by supermarkets (43%), convenience stores (1%), and other retailers [19,38]. Efforts should focus on increasing the accessibility and affordability of nutritious foods such as fruits and vegetables, particularly in grocery stores. Additionally, in this digital age, an online delivery platform that leverages the proximity of these stores to households in Putrajaya can promote the purchase and delivery of fresh produce to homes and food premises. Close collaboration with farmers as well as meat, poultry, and seafood suppliers can further boost engagement and coordination of such arrangements.

On the policy front, implementing a tax on less nutritious foods, combined with a 20% reduction in the price of healthy food options, could shift consumer preferences towards healthier food choices [39]. Revenues from these taxes can be directed to support small-scale local farmers in competing with large-scale agricultural monopolies. On a larger scale, subsidising manufacturers to produce healthier food items may encourage consumers to choose healthier food when the cost of basic food items bread, milk, and frozen food is adjusted by the manufacturers.

Moreover, expanding the Ministry of Health’s “*Kafetaria Sihat*” certification to more eateries may help foster healthier dining options [40]. Furthermore, there is a need to strengthen the promotion of healthy food options through innovative marketing approaches, such as food samples, in-store urban farming, recipe showcases, and healthy food festivals. These strategies can inspire individuals to incorporate more nutritious ingredients into their diets. Nutritional information on the menu may also enhance customers’ perceptions of the food quality [41]. As more consumers are now acquiring food information online, nutrition awareness and healthy eating promotion must also leverage digital channels such as social media and podcasts.

Besides that, as with many developing countries, Malaysians have a reputable norm of long working hours. The best strategy to improve food quality and encourage home-cooked meals is to promote work-life balance. Besides, it will be helpful to the cause of healthy eating if pre-made, well-balanced nutritious dishes can be made tasty and affordable. Fruits and vegetables should also be promoted as convenient alternatives to food in the face of urban lifestyles [42]. Initiatives to reformulate products to increase vegetable and fibre content while also reducing calories, fats, sugar, and sodium should be encouraged.

### 4.2 Limitation

This study only assessed food stores in the locality of Putrajaya. However, some residents might have chosen to dine and purchase groceries from other locations rather than exclusively visiting the store in the closest proximity to them. Besides, brand-switching is a common occurrence in view of the high competitiveness in the food and beverages businesses. In addition, street markets such as weekly farmer markets or night markets, which may offer products such as fresh fruits and vegetables, were not assessed in the study. Last but not least, the tools used in this study did not take into account the portion size and the amount of calories, sugar, and salt in the meal. Future studies should assess food outlets beyond Putrajaya to capture actual purchasing behaviours and brand-switching. Street markets and other informal food sources should also be included. Additionally, incorporating portion size and nutritional content such as calories, fats, sugar, and salt, would provide a more comprehensive evaluation of food healthfulness.

## 5.0 CONCLUSION

The disparity in food healthfulness in Putrajaya highlights the urgent need to improve the local food environment. A comprehensive strategy that enhances the availability, accessibility, and affordability of nutritious foods, alongside advocating for healthy eating is essential. Supportive policies and legislation can strengthen these efforts, thus creating an environment that encourages healthier dietary practices. Promoting such a food environment is a critical step toward disease prevention and improved population health.

## Data Availability

The dataset is available via Google Drive at the following link: https://docs.google.com/spreadsheets/d/1mMBwPyYQigI7nnU78ONT3KqPuOMGwyJM/edit?usp=sharing&ouid=100276414262675383274&rtpof=true&sd=true

https://docs.google.com/spreadsheets/d/1mMBwPyYQigI7nnU78ONT3KqPuOMGwyJM/edit?usp=drive_link&ouid=100276414262675383274&rtpof=true&sd=true

## Acknowledgments

The authors sincerely thank colleagues at the Faculty of Medicine and Health Sciences, Universiti Putra Malaysia (UPM), for their valuable input. We are also grateful to the business owners in Putrajaya who were involved in the study, and to the Putrajaya Corporation and the Department of Survey and Mapping Malaysia (JUPEM) for providing access to the spatial datasets used in this study.

